# Changes in pneumococcal deaths in the United States following the COVID-19 pandemic

**DOI:** 10.1101/2025.02.24.25322782

**Authors:** Lianhan Shang, Stephanie Perniciaro, Daniel M Weinberger

## Abstract

**Background:** Although changes in the rates of pneumococcal cases during the COVID-19 pandemic have been extensively described, changes in rates of death due to pneumococcus during this period are not well understood.

**Methods:** We obtained vital statistics data for the United States (National Center for Health Statistics), including age, sex, race/ethnicity, cause of death (ICD-10), 2014-2022. Generalized linear models were fit to the period from January 2014-February 2020 and extrapolated to March 2020-December 2022 to generate an expected number of pneumococcal deaths and a 95% prediction interval. We used a lasso regression model to identify clinical and demographic factors most strongly associated with pneumococcal deaths during the pandemic period as compared with a pre-pandemic baseline.

**Results:** For most of 2020, pneumococcal deaths were not notably different from the pre-pandemic period and largely followed the typical seasonal pattern. However, at the end of 2020 and early 2021, when pneumococcal deaths would typically peak, the rates of death remained lower than normal and stayed lower than expected in the spring of 2021. Starting around mid-2021, there was a notable spike above baseline that coincided with the Delta wave of COVID-19. The 2021 winter – 2022 spring peak followed the pre-pandemic trend. Some of the changes could be attributed to changes in the seasonality of respiratory viruses that interact with pneumococcus. The prevalence of certain risk factors among pneumococcal deaths elevated following the pandemic, including obesity (OR = 1.40), diabetes mellitus without complication (OR = 1.39) and heart failure (OR = 1.31).

**Conclusions:** The COVID-19 pandemic significantly disrupted pneumococcal mortality patterns in ways that were distinct from the changes described in clinical cases of invasive pneumococcal disease.

**Key points:** COVID-19 pandemic significantly disrupted pneumococcal mortality patterns. Respiratory viruses are important trigger for pneumococcal diseases.

## INTRODUCTION

Pneumococcus is an important bacterial pathogen that causes a range of conditions including pneumonia, otitis media and invasive pneumococcal disease (IPD) [1]. IPD is defined based on the isolation of pneumococcus from normally sterile sites (blood, cerebrospinal fluid). During the first year of the COVID-19 pandemic, reported rates of IPD declined in many countries[1–3]. It was initially thought that the declines in rates of IPD were caused by reduced transmission of pneumococcus related to non-pharmaceutical interventions [4]. However, subsequent studies have suggested that the prevalence of carriage of pneumococcus was relatively stable during the pandemic [5–8], with conflicting evidence about whether the average density of colonization declined [6, 8].

There are alternative explanations for the decline in rates of IPD. First, there is evidence that the risk of IPD greatly increases following infection with certain respiratory viruses, such as influenza and respiratory syncytial virus (RSV). Many of these seasonal viruses disappeared during the first year of the pandemic, removing a potential trigger for developing IPD. Supporting this, declines and subsequent resurgence of rates of IPD correlated with the activity of respiratory viruses, including influenza, respiratory syncytial virus (RSV), and human metapneumovirus (hMPV)[9]. It is also possible that some of the decline in IPD during the initial phase of the pandemic was due to changes in diagnostic practices[10]. However, the decline in rates of IPD was seen in many countries [2], each with distinct epidemiological settings and testing guidelines.

While the decline in the rate of IPD has been extensively described, there has been little consideration for how the pandemic might have influenced rates of death due to pneumococcal disease. It might be expected that if cases of pneumococcal disease decline, deaths would also decline. Trends in rates of death could differ from trends in reported cases if the population of people dying differs in important ways (exposure to the bacteria, susceptibility to severe infection) from the broader population of people getting sick with pneumococcal disease. Likewise, if there are changes in the use of clinical diagnostics over time, this could affect some outcomes more than others. In this study, we evaluated changes in rates of deaths recorded as being related to pneumococcus during the post-pandemic period using nationwide vital statistics data from the United States and evaluated changes in the characteristics of these deaths.

## METHODS

### Data sources

The data for this study were obtained from the National Center for Health Statistics (NCHS). These individual-level data are publicly available and can be downloaded from the NCHS website [11]. These vital statistics data have detailed information on the deceased individual including age (years), sex, race/ethnicity, and the underlying and contributing causes of death (coded using the International Classification of Diseases, 10^th^ revision [ICD-10]). We included deaths of individuals ≥ 25 years in the NCHS database. The weekly number of influenza cases was obtained from CDC Fluview [12]. The weekly number of RSV cases was obtained from The National Respiratory and Enteric Virus Surveillance System (NREVSS) [13]. The population data were from CDC WONDER [14].

### Definition of outcomes

The primary outcome of interest was death with pneumococcal disease. Among deaths recorded in the database, we defined pneumococcal deaths as those with pneumococcal-specific ICD-10 codes (A40.3, J13, B95.3, or G00.1) anywhere in the death record. Other outcomes were death with pneumococcal pneumonia, and death with non-respiratory IPD. We defined death with pneumococcal pneumonia as the presence of either a specific pneumococcal pneumonia code (J13) or both a pneumococcal code (A40.3, J13, B95.3, or G00.1) and any pneumonia code (J12-J18). IPD was defined as cases with both a pneumococcal code and either sepsis (A40, A41.9) or meningitis (G00) codes. Deaths considered to be non-respiratory IPD were those with the IPD codes but without pneumococcal pneumonia codes.

### Statistical analysis

For the main analyses on all deaths with pneumococcal disease codes, we compared the observed and expected number of deaths during each month of the pandemic period (March 2020-December 2022). The expected number of deaths, and 95% prediction intervals, were estimated using a generalized linear regression model that adjusts for underlying trends and seasonality, as described previously[15]. The model was fit to the pre-pandemic period (January 2014-February 2020) and then extrapolated to the pandemic period (March 2020-December 2022). We calculated the rate ratio (RR) and 95% CI as the ratio of observed to expected cases during each month. We conducted sensitivity analyses further adjusting for the monthly standardized influenza or RSV cases. The weekly data of influenza and RSV were standardized by calculating the ratio of positive cases to the 52-week centered moving average of testing volume (multiplied by 100) to account for temporal variations in testing intensity. The monthly standardized cases were the average of the weekly data using weights proportional to the number of days each week contributed to that month. In addition, we conducted subgroup analyses for age group, education and race/ethnicity categories. The secondary analyses were conducted on deaths with pneumococcal pneumonia codes and deaths with non-respiratory IPD codes.

To investigate the change of underlying diseases/demographic characteristics associated with pneumococcal death during the pandemic, we performed a lasso regression on Clinical Classifications Software Refined (CCSR) categories (whose frequency is > 100 in the database, and is not COVID-19) and demographic characteristics comparing the data during the pandemic with pre-pandemic data. The CCSR categories were defined based on the ICD-10 codes on the death certificates.

### Availability of code and data

All analysis were run on R 4.3.1. The codes are available from https://github.com/PneumococcalCapsules/pneumococcal_mortality. Data for all analyses are available from https://zenodo.org/records/12808102.

## RESULTS

### Characteristics of pneumococcal deaths

There were 8590 deaths with pneumococcal disease codes among people ≥ 25 years of age in U.S. from 2014 to 2022 (Table 1). Roughly 63% of the deaths occurred in adults age ≥65 years. Females accounted for 47% of the deaths. The White population accounted for more than 70% of the deaths. More than 60% of the deaths happened among people with high school and lower education. There were 6068 deaths with pneumococcal pneumonia codes and 2522 deaths with non-respiratory IPD. The distribution of age, sex, race/ethnicity and education for deaths with pneumococcal pneumonia and non-respiratory IPD was similar to overall pneumococcal deaths (Table 1).

**Table 1.**
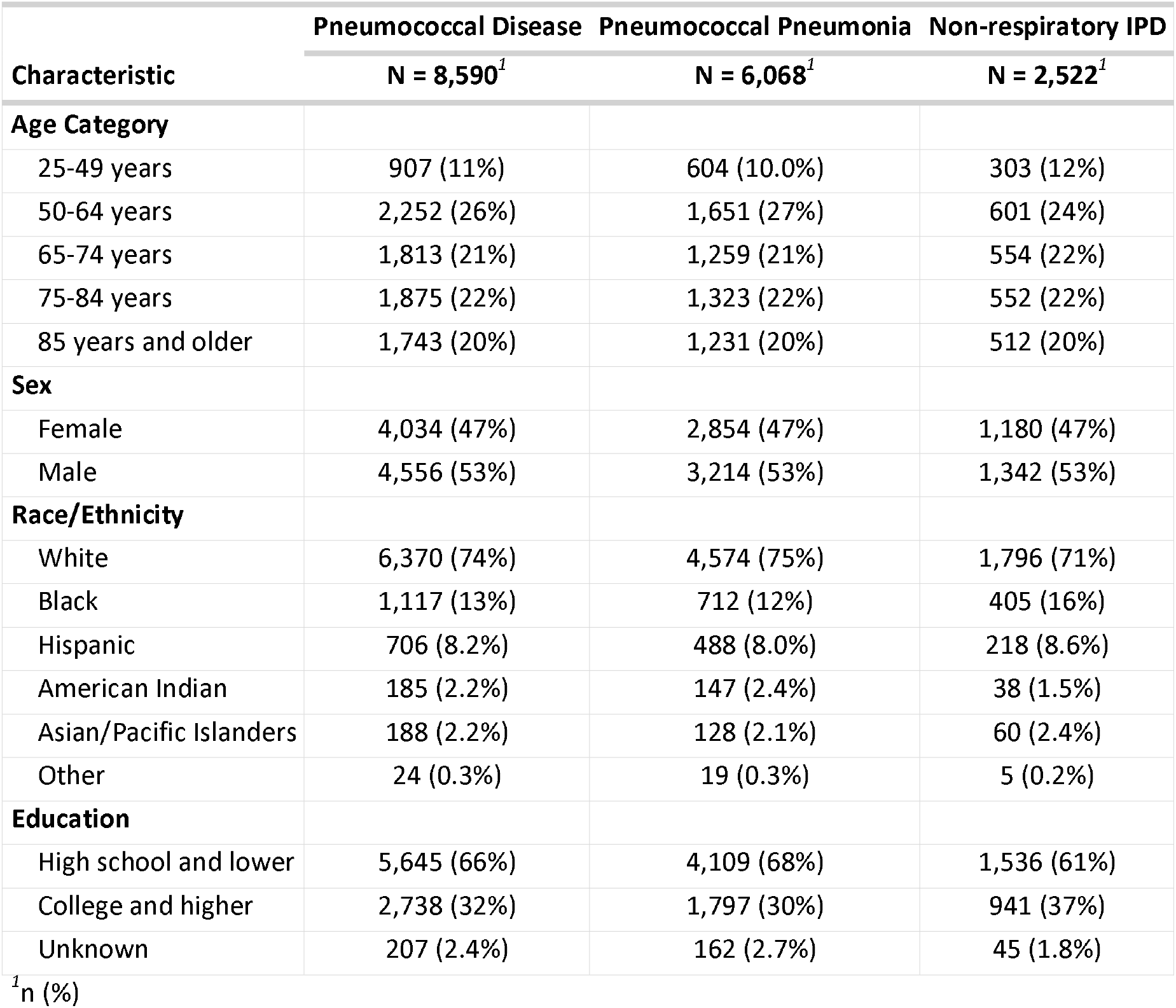
Characteristics of pneumococcal deaths.

Rates of death with pneumococcus varied by race/ethnicity, with the highest rates among American Indian populations and the lowest rates among individuals identified as Asian/Pacific Islanders, those identified as Hispanic (Figure S1). Black individuals had higher rates than White individuals for the youngest age group and through young and middle-aged adult age groups, but these differences largely disappeared for those over the age of 75 years.

### Changes in rates of death with pneumococcal codes during the 2020-2022 COVID-19 pandemic

During the pre-pandemic period, rates of death with pneumococcal disease were highly seasonal, with peaks commonly occurring during late-winter and early-spring (Figure 1A). For most of 2020, including the early months of the COVID-19 pandemic when the US was experiencing lockdowns, the reported deaths were not notably different from the pre-pandemic period and largely followed the typical seasonal pattern. However, at the end of 2020 and early 2021, when pneumococcal deaths would typically peak, the rates of death remained lower than normal and stayed lower than expected in the spring of 2021 (Figure 1A, Figure 1D). Starting around mid-2021, there was a notable spike above baseline that coincided with the Delta wave of COVID-19 (Figure 1A, Figure 1D). The 2021 – 2022 winter peak followed the pre-pandemic trend. After adjustment for the activity of influenza, the difference between predicted rates and observed rates of pneumococcal deaths became much smaller in the 2020– 2021 season (Figure 1B, Figure 1E). However, the RR was still high in the peak which coincided with the Delta wave. Adjusting for RSV activity produced a similar patterns to adjusting for influenza activity (Figure 1C, Figure 1F).

**Figure 1.**
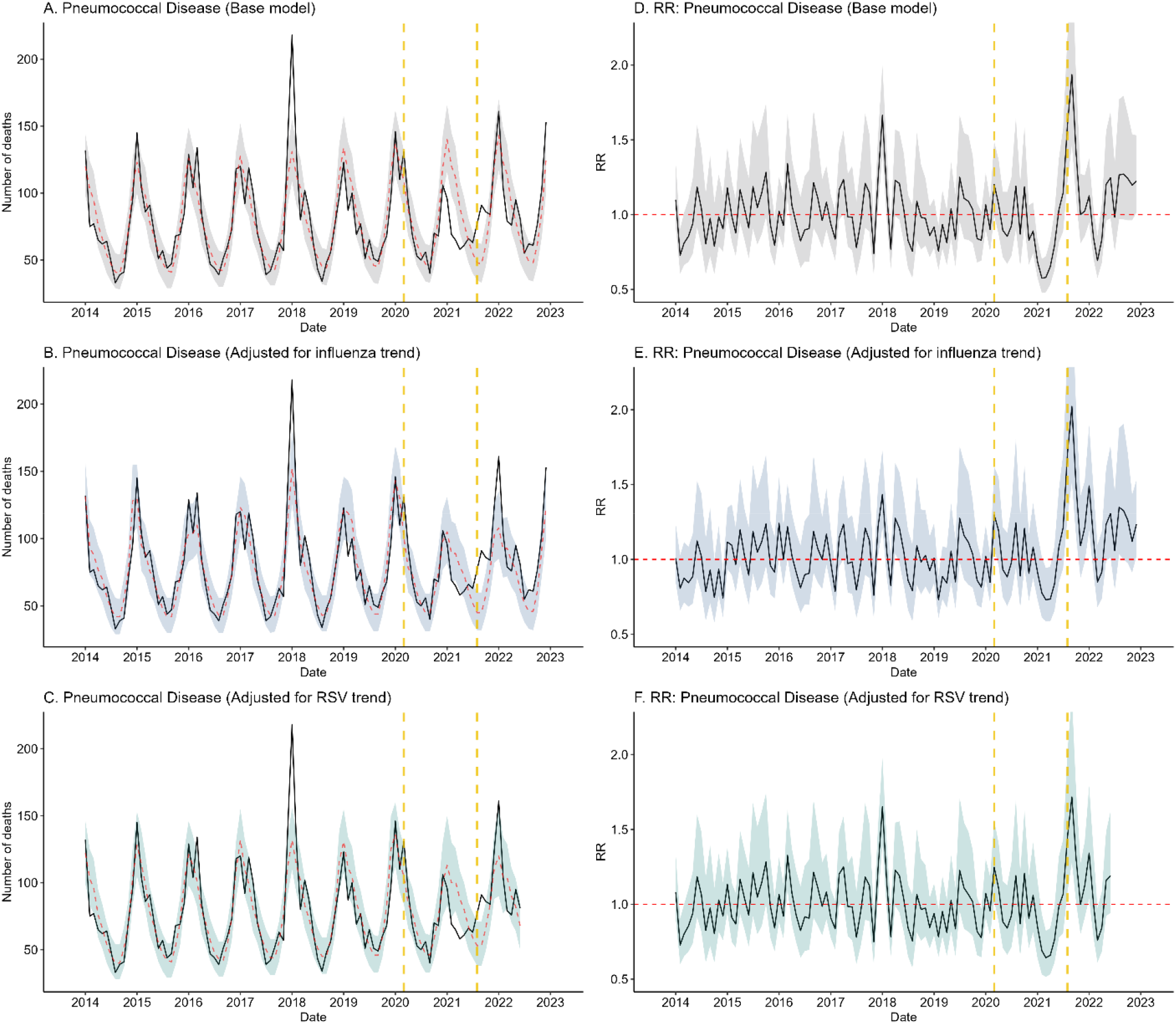
Time series for deaths and RR of pneumococcal disease

In subgroup analysis for age, people younger than 75 years old had comparatively higher RR in the death peak coinciding with Delta wave (Figure 2A). People with different education backgrounds showed similar changes from baseline (Figure 2B). Among the race/ethnicity groups, non-White groups had larger increases above the baseline than White people during the Delta wave (Figure 2C). The subgroup analyses using the model adjusting for influenza/RSV trend showed similar results (Figure S2, Figure S3)

**Figure 2.**
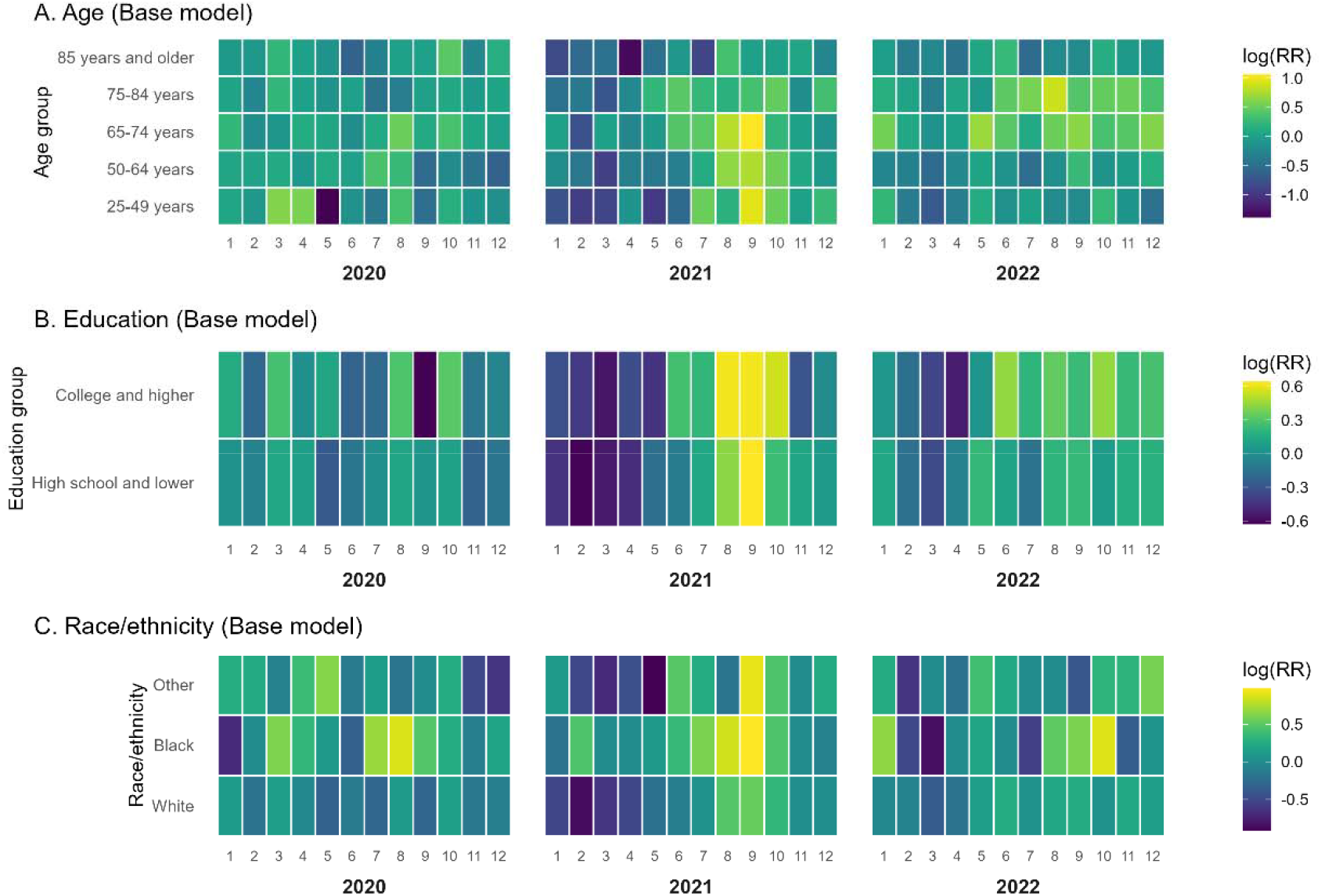
Heatmap for subgroup analyses (pneumococcal disease)

**Figure 3.**
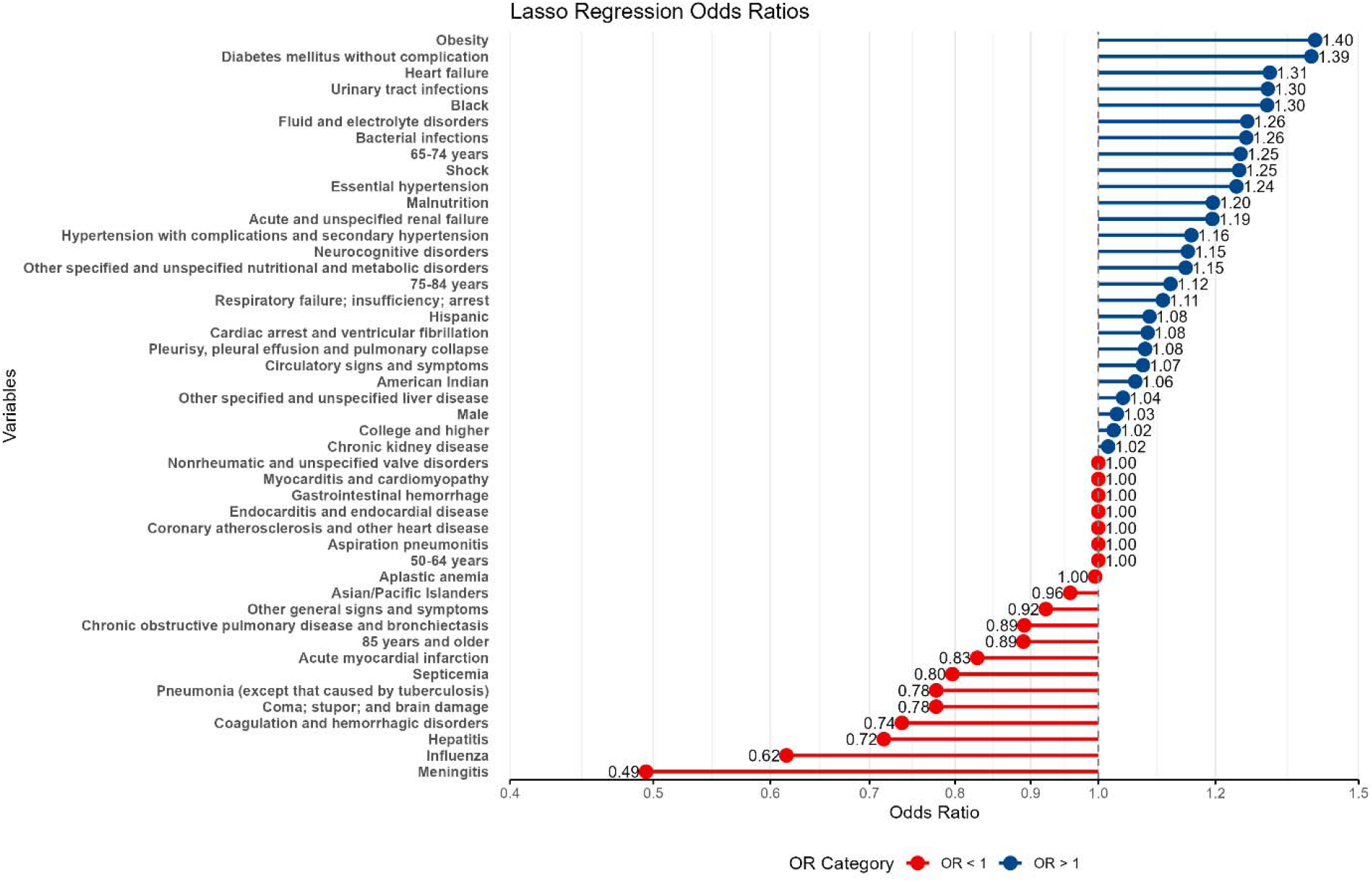
Lollipop plot for lasso regression

### Changes in rates of death with pneumococcal pneumonia codes and non-respiratory IPD codes during the 2020-2022 COVID-19 pandemic

Deaths with pneumococcal pneumonia and non-respiratory IPD also followed the seasonal pattern in the pre-pandemic period (Figure S4A, Figure S5A). Their peak in the 2020-2021 season was also much lower than predicted (Figure S4A & D, Figure S5A & D) and showed a peak which coincided with Delta wave. Adjusting for influenza and RSV trend partially mitigated the differences between predicted deaths and observed deaths in the 2020 winter – 2021 spring season but did not affect the peak coinciding with Delta wave (Figure S4 BC & EF, Figure S5 BC & EF).

### Changes in the frequency of comorbid conditions among pneumococcal deaths

We evaluated shifts in the prevalence of clinical and demographic factors associated with pneumococcal deaths during the pandemic period as compared with a pre-pandemic baseline and observed a notable shift in underlying conditions. The CCSR categories with highest ORs were obesity (OR = 1.40), diabetes mellitus without complication (OR = 1.39) and heart failure (OR = 1.31), indicating that these conditions were more commonly identified among pneumococcal deaths during the pandemic period. In contrast, certain conditions such as meningitis (OR = 0.49), influenza (OR = 0.62), hepatitis (OR = 0.72), and coagulation and hemorrhagic disorders (OR = 0.74) were associated with lower odds to co-occur with pneumococcal deaths relative to the pre-pandemic era. Black race (OR = 1.30) and the age group 65-74 years (OR = 1.25) were more commonly identified among pneumococcal deaths during the pandemic.

## DISCUSSION

Our analysis revealed temporal patterns in pneumococcal deaths during the COVID-19 pandemic that were distinct for those described for clinical cases of invasive pneumococcal disease. In the initial months of the pandemic, pneumococcal mortality patterns remained largely unchanged, unlike the rapid drops reported for IPD. However, a significant deviation from historical patterns emerged during the winter of 2020-2021, when deaths were notably lower expected. An unusual peak in pneumococcal deaths coincided with the Delta wave of COVID-19 in the autumn of 2021. The subsequent winter season (2021-2022) showed a return to typical seasonal patterns. Adjusting for influenza or RSV trends reduced the difference between predicted and observed deaths during the winter of 2020-2021. However, neither adjustment explained the unusual peak during the Delta wave. Both deaths associated with pneumococcal pneumonia and non-respiratory IPD showed temporal patterns similar to overall pneumococcal deaths.

Although prior studies demonstrated a drop in pneumococcal disease and IPD incidence following the onset of the COVID-19 pandemic [1–3, 16, 17], our findings indicated that pneumococcal-related deaths did not decrease substantially until late 2020. This is consistent with data from the CDC’s active bacterial core surveillance system, which reported a steeper decline in cases of IPD than deaths due to IPD. The discrepancy between changing pattern of pneumococcal cases and pneumococcal-related deaths may be due to the health-seeking behavior and diagnostic practices during the pandemic. Public health interventions such as social distancing, masking and lockdown were rapidly implemented after the outbreak of SARS-CoV-2 infections [18, 19] but largely did not affect colonization rates in the community [5–8]. In addition, medical resources were prioritized towards COVID-19 patients in the first several months of COVID-19 pandemic. Consequently, individuals with mild to moderate pneumococcal disease may have avoided or delayed seeking care [20, 21]. Even those who did present to hospitals might lack access to diagnostic testing, thereby receiving only empirical treatment and potentially reducing the recorded incidence of pneumococcal disease and IPD. In contrast, severe cases that were more likely to lead to deaths might be managed with the same level of urgency and standard of care as before the pandemic, and would have a had higher chance to be properly recorded in the medical documents, which might explain why the mortality rates do not drop as sharply as the reported case numbers.

In late 2020 and early 2021, observed pneumococcal deaths were much lower than predicted. This trend was consistent with several studies reporting significant reductions in pneumococcal hospitalizations and overall case numbers [1–3, 16, 17, 22]. Despite the decrease in clinical disease, previous evidence from different regions of the world showed that pneumococcal carriage was relatively stable in the population [5–7, 23]. One possible explanation of the decline in pneumococcal death is that public health interventions disrupted the transmission of respiratory viruses that can facilitate pneumococcal invasion and exacerbate disease severity [24]. Respiratory viruses, such as influenza and RSV, may cause epithelial damage, facilitate dysfunction of lung physiology, up-regulate pneumococcal receptors and alternate immune responses towards the proinflammatory direction, which altogether increases hosts’ vulnerability to adherence and invasion of pneumococcus [25–27]. Epidemiological evidence suggested that influenza and RSV infection was associated with higher risk of pneumococcal diseases [9, 24, 28, 29]. In our study, adjusting for the trend of influenza and RSV greatly mitigated the gap between the expected and observed pneumococcal mortality, echoing the role of respiratory viruses in the pathogenesis of pneumococcal disease.

Another important finding of our study was the unusual peak of pneumococcal deaths in the summer of 2021. Adjustment for influenza and RSV could not account for the peak, while the peak coincided with the Delta wave of COVID-19. Therefore, a potential explanation for the peak is that SARS-CoV-2 may also serve as a trigger for pneumococcal disease, similar to influenza and RSV. SARS-CoV-2 infection could also disrupt physical barriers and cause dysregulated immune system, which increase the risk of pneumococcal disease [30–32]. In subgroup analysis, Black and other racial/ethnic groups experienced disproportionately higher pneumococcal mortality during the Delta wave, reflecting persistent healthcare inequities that demand policy interventions [33, 34].

When comparing pre- and post-pandemic pneumococcal deaths using CCSR categories, obesity had the highest odds ratio among all comorbidities. This finding builds on existing evidence that obesity can compromise immune responses, impair respiratory mechanics, and exacerbate inflammatory processes, all of which predispose individuals to severe outcomes from bacterial pneumonia [35–37]. The impact of obesity may have been amplified during the pandemic as timely healthcare may have been less accessible and physical activity was reduced during lockdowns [38, 39]. Thus, the diagnosis and treatment of pneumococcal infections might be delayed during the pandemic. While prior research has primarily focused on obesity as a key risk factor for severe respiratory infections, our results suggest that the interaction between obesity and pandemic-related factors is also important in determining disease profile.

The primary limitation of our study was the reliance on ICD codes to identify pneumococcal deaths, which may lead to misclassification. Some deaths labeled with pneumococcal codes may have been caused by other organisms or may not have received confirmatory testing. Additionally some deaths that should have been attributed to pneumococcus may have been coded differently: in an audit of coding practices from 2018-2022 at a large hospital, around half of cases of hospitalized pneumonia with a positive pneumococcal urine antigen test were not coded as J13 pneumococcal pneumonia (most of these were coded as J15.4, pneumonia due to other streptococci and J18.9, pneumonia due to unspecified organism). Besides, the shifts in diagnostic protocols and healthcare-seeking behavior during the pandemic may have introduced biases which we could not account for in the model.

In summary, the COVID-19 pandemic significantly disrupted pneumococcal mortality patterns, as reflected by a marked reduction during the initial winter wave that aligned with changes in respiratory virus activity. Besides, there was a notable surge of pneumococcal death coinciding with Delta wave of COVID-19 which could not be explained by influenza or RSV, suggesting the potential role of SARS-CoV-2 as a trigger for pneumococcal disease. Our findings highlighted the influence of respiratory viruses on the development and severity of pneumococcal disease.

## Funding

This study was supported by a grant from the Merck Investigator Studies Program to Yale University. The funder had no role in the design, conduct or reporting of the study.

## Disclosures

DMW has received consulting fees from Pfizer, Merck, and GSK for work unrelated to this manuscript and has been principal investigator on research grants from Pfizer and Merck to Yale University.

## Data Availability

Data for all analyses are available from https://zenodo.org/records/12808102.

## Notes

### Author Declarations

National Center for Health Statistics (NCHS) website

