## Supplementary material for "Changes in pneumococcal deaths in the United States following the COVID-19 pandemic": Figure S1

Figure S1 Rate of death with pneumococcus by age and race/ethnicity

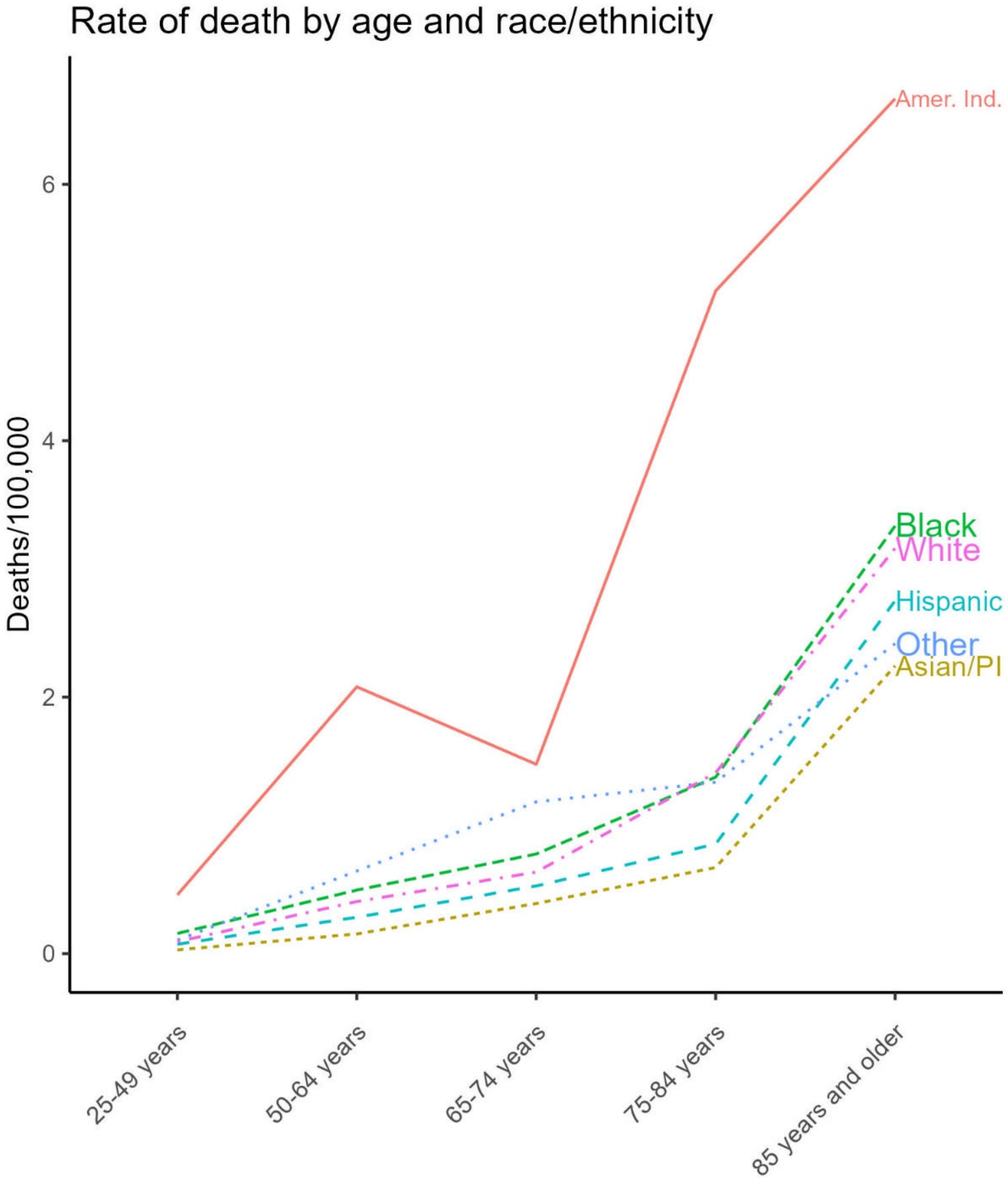

Figure S2 Heatmap for subgroup analyses (pneumococcal disease, adjusted for influenza trend)

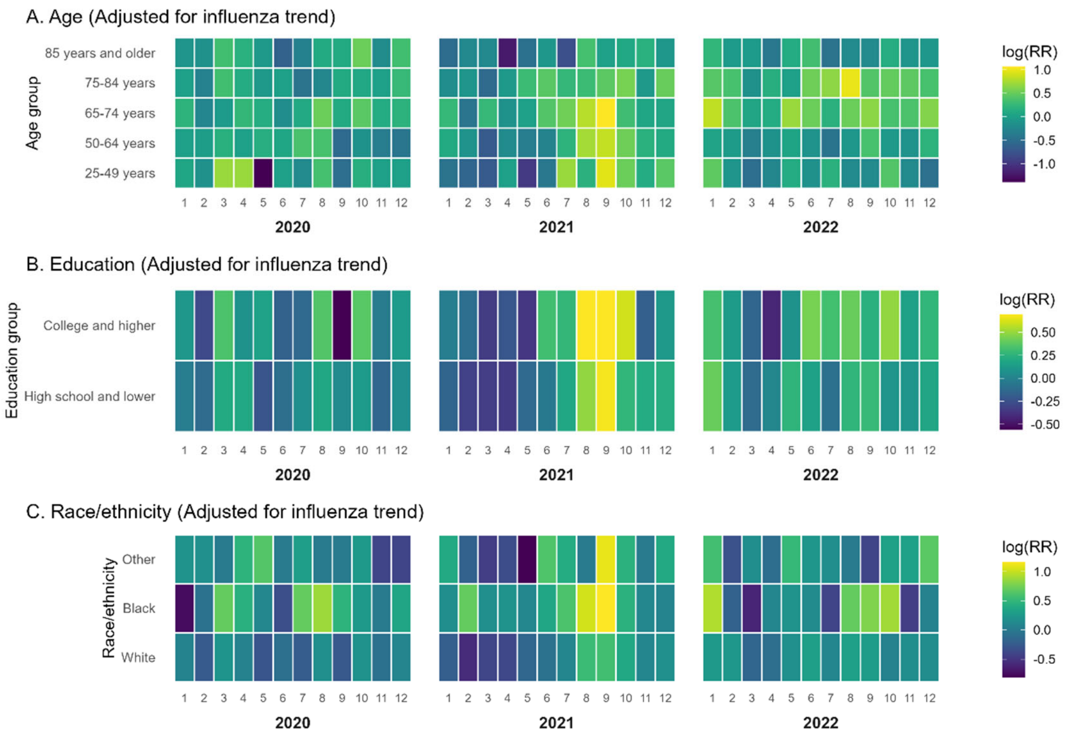

Figure S3 Heatmap for subgroup analyses (pneumococcal disease, adjusted for RSV trend)

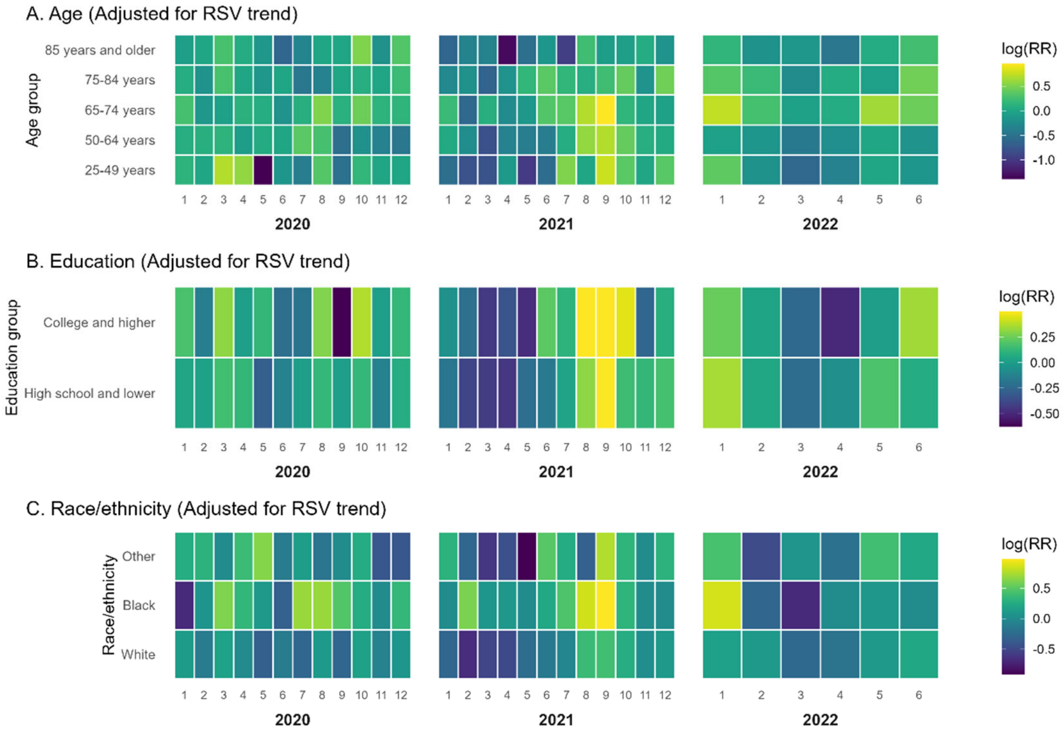

Figure S4 Time series for deaths and RR of pneumococcal pneumonia

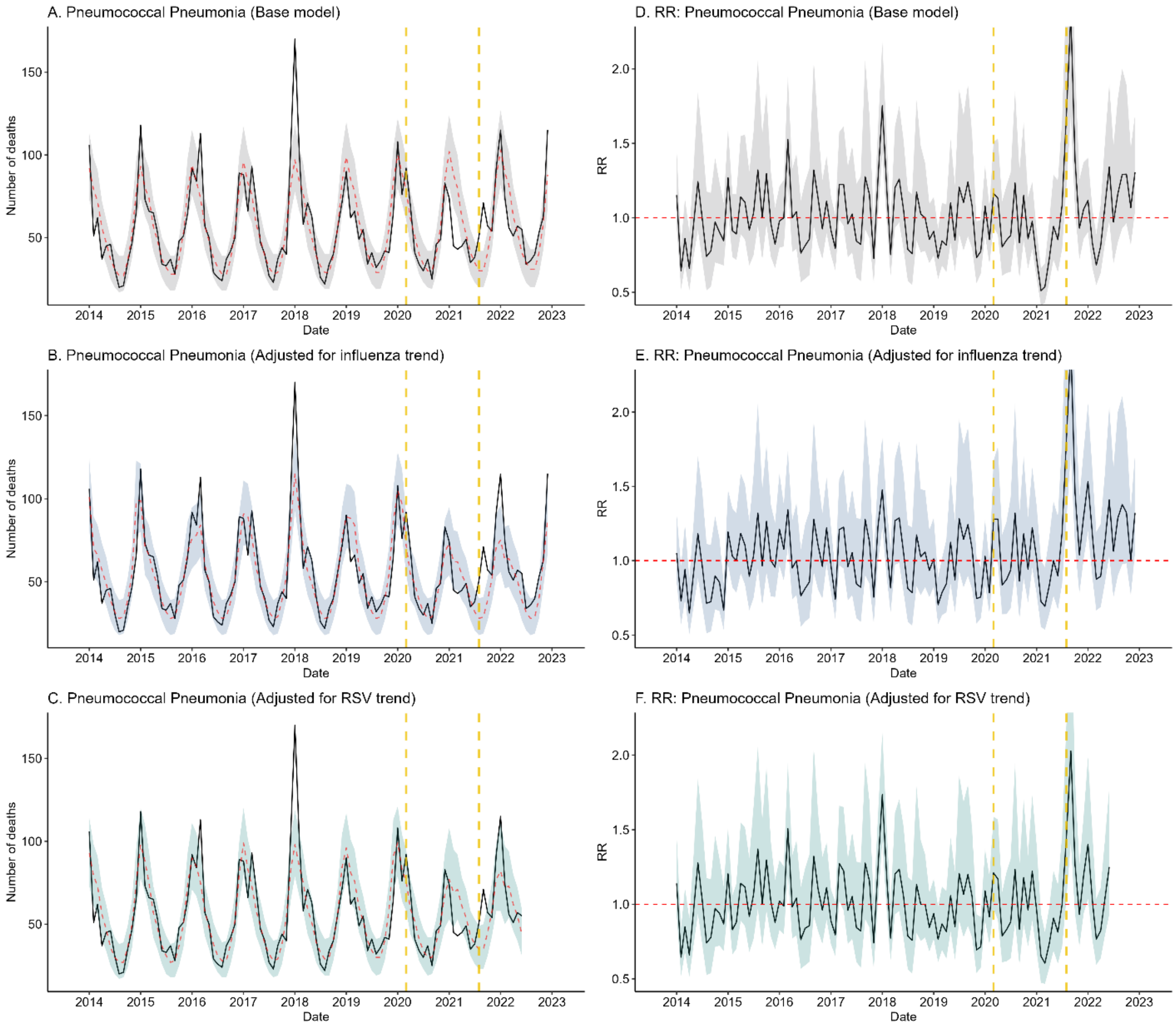

### Figure S5 Time series for deaths and RR of non-respiratory IPD

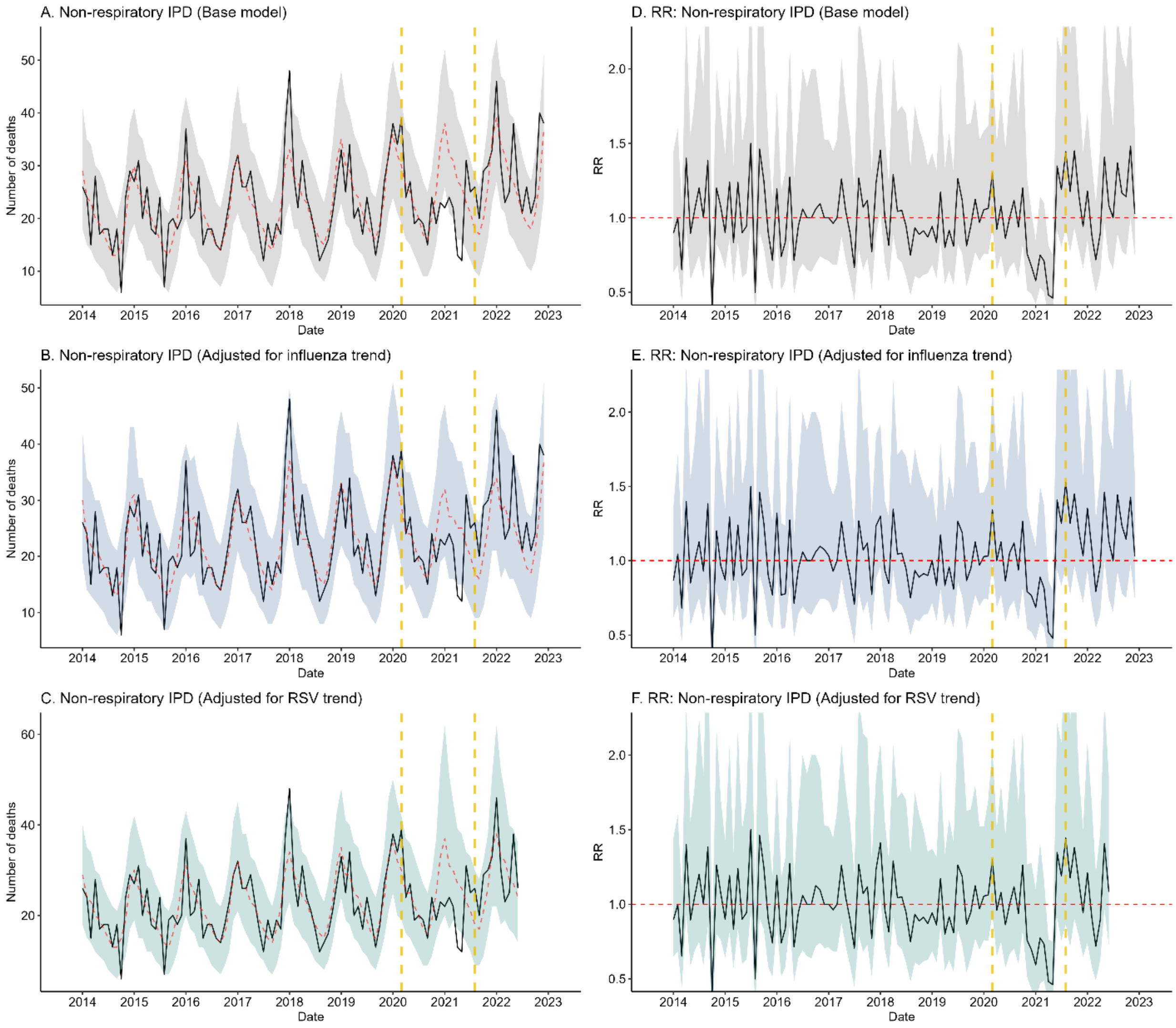
